# A Bayesian Logistic Growth Model for the Spread of COVID-19 in New York

**DOI:** 10.1101/2020.04.05.20054577

**Authors:** Svetoslav Bliznashki

## Abstract

We use Bayesian Estimation for the logistic growth model in order to estimate the spread of the coronavirus epidemic in the state of New York. Models weighting all data points equally as well as models with normal error structure prove inadequate to model the process accurately. On the other hand, a model with larger weights for more recent data points and with t-distributed errors seems reasonably capable of making at least short term predictions.

## 1. Introduction

The logistic growth model is frequently used in order to model the spread of viral diseases and of covid-19 in particular (e.g. Batista, 2020; Wu et al., 2020). The differential equation is given in (1):

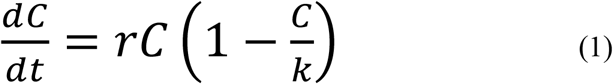

where C is the cumulative number of infected individuals, r is the infection rate, and K is the upper asymptote (i.e. the upper limit of individuals infected during the epidemic). Unlike other models, like SIR, Eq. 1 has an explicit analytical solution:

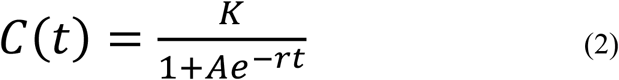

where A=(K-C_0_)/C_0_ and C_0_ is the initial number of infectees.

The parameters of Eq. 2 can easily be estimated via Least Squares (LS) but in this note we use a Bayesian approach which allows us to make use of explicit posterior distributions in order to make probabilistic predictions.

We apply the above model to the state of New York which represents a relatively geographically homogenous population with sufficient data points in order build a reliable model which is not affected by different trends present in different regions.

## 2. Simulation 1

We begin with a simple model which estimates the parameters of Eq. 2 based on the assumption of normally distributed homoscedastic errors. We use the data for 28 consecutive days of the epidemic beginning with the 4^th^ of March (11 infectees) and ending with the 31^st^ of March (75832 infectees)^1^.

Prior to estimation we standardized our data by dividing all data points by 70000 in order to avoid numerical problems; after the posteriors were obtained we back-transformed the results in their original scale.

We assumed that the errors are normally distributed with mean equal to 0 and standard deviation (σ) estimated by the model.

We used the blockwise Random Walk Metropolis algorithm^2^ in order to sample from the joint posterior distribution of the four parameters of the model (K, A, r, and σ). The proposal distribution was multivariate normal with scaled variance-covariance matrix estimated on the basis of pilot runs. Uninformative improper uniform priors ranging from 0 to + ∞ were employed for all parameters in the model. A pilot chain showed an acceptance rate within the optimal range of 23% (e.g. Chib & Greenberg, 1995). The final simulation used 20 million iterations and heavy thinning (each 200^th^ sample was retained). The traceplots and autocorrelation functions indicated excellent convergence. A histogram and a traceplot for the K parameter are shown in Fig. 1.

**Figure 1.**
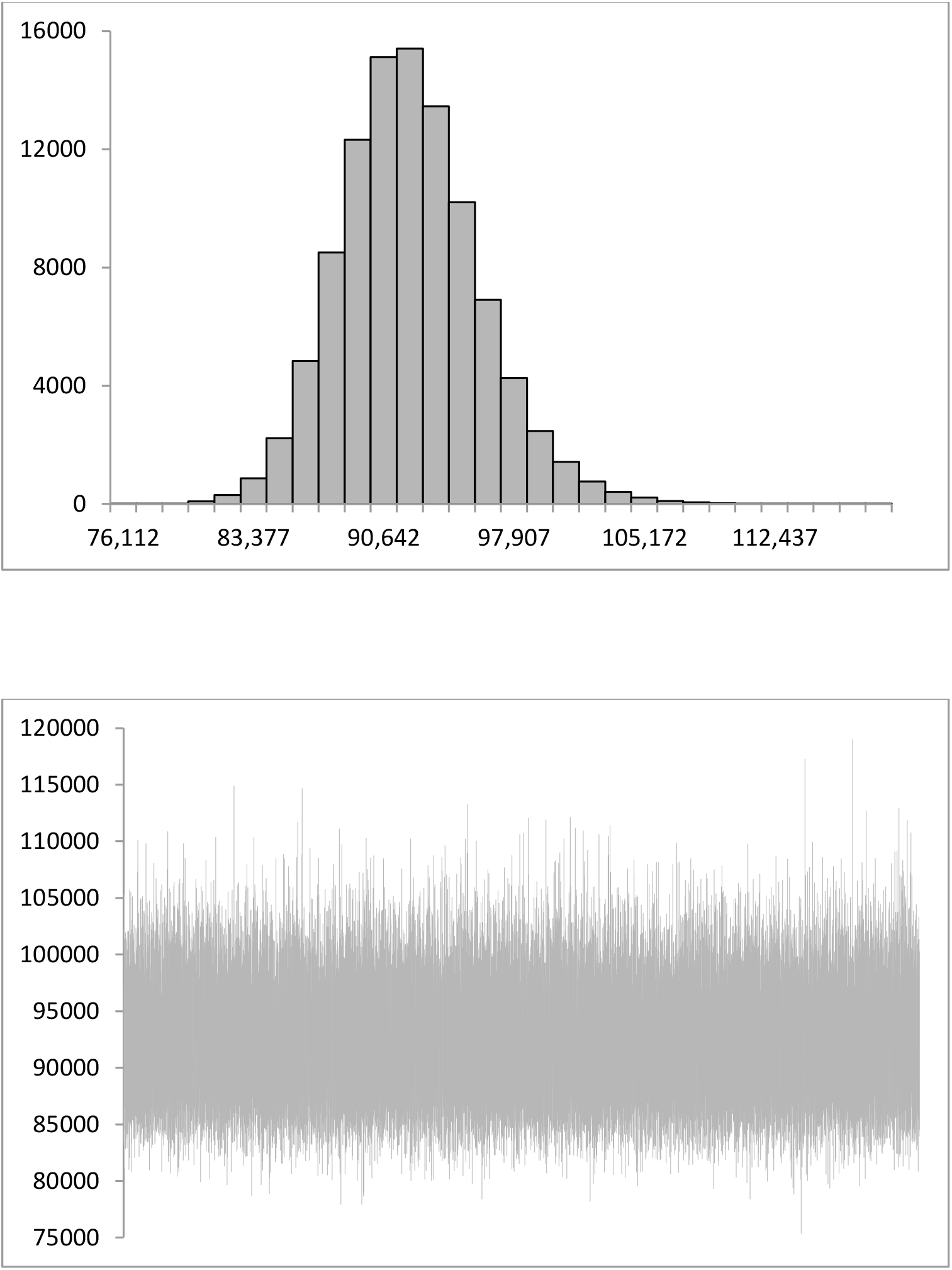
A histogram (top) and a traceplot (bottom) for the posterior for the K parameter from Eq. 2 for our first model.

Table 1. Means, medians, standard deviations (SD), and 95% HDIs for the parameters of Eq. 2 obtained from the posterior of our first model.

**Table 1.** Shows the mean (Expected Aposteriori - EAP) and median estimates for the 4 parameters as well as their standard deviations and 95% Highest Density Intervals (HDI).

|  | mean | median | SD | 2.50% | 97.50% |
| --- | --- | --- | --- | --- | --- |
| K | 92114.87 | 91903.83 | 3904.86 | 84636.23 | 99978.49 |
| A | 2674.25 | 2564.55 | 707.51 | 1473.55 | 4069.47 |
| r | 0.34 | 0.34 | 0.01 | 0.31 | 0.37 |
| $\sigma$ | 0.02 | 0.02 | 0.003 | 0.01 | 0.02 |

<sup>1</sup> The data is publicly available via the github repository: https://github.com/nytimes/covid-19-data/blob/master/us-states.csv
<sup>2</sup> The posterior chains showed high correlations between the parameters (ranging from -0.87 to +0.96) which renders algorithms like the Gibbs Sampler and/or componentwise Metropolis highly inefficient for the task at hand.

Note that in Table 1 and in subsequent tables of this type the σ parameter and its posterior SD are given in the standardized scale (the original data was divided by 70000 as explained above) in order to avoid clutter. The same is true for the scale parameter described in Section 4 (Table 5). The K parameter is always given in its original (unstandardized) form since it is of primary interest.

**Table 5.** Means, medians, standard deviations (SD), and 95% HDIs for the parameters of Eq. 2 obtained from the posterior of our last model.

|  | mean | median | SD | 2.50% | 97.50% |
| --- | --- | --- | --- | --- | --- |
| K | 169235.12 | 170739.00 | 19085.84 | 128757.85 | 202630.50 |
| A | 293.33 | 276.99 | 51.09 | 232.13 | 403.94 |
| r | 0.20 | 0.20 | 0.01 | 0.18 | 0.23 |
| scale | 0.01 | 0.01 | 0.002 | 0.002 | 0.01 |
| df | 5.79 | 2.38 | 10.32 | 1.03 | 23.30 |

<sup>4</sup> The lognormal distribution is not symmetric and hence we use the actual Metropolis-Hastings acceptance probability (e.g. Chib & Greenberg, 1995) during the step sampling from the df posterior.

For comparison, the estimates obtained from LS estimation (via the Matlab’s cftool) are the following: K=92470 (95% CI=[85120; 99890]), A=2424 (95% CI=[1247; 3601]), and r=0.34 (95% CI=[0.31; 0.37]). We see that the estimates are very similar to the posterior EAPs reported above which is to be expected given the uninformative nature of our priors. Still, the Bayesian analysis gives slightly wider intervals for the estimates which as we’ll see below is a positive.

Fig. 2 below shows the observed and the fitted estimates for the cumulative number of infectees in New York.

**Figure 2.**
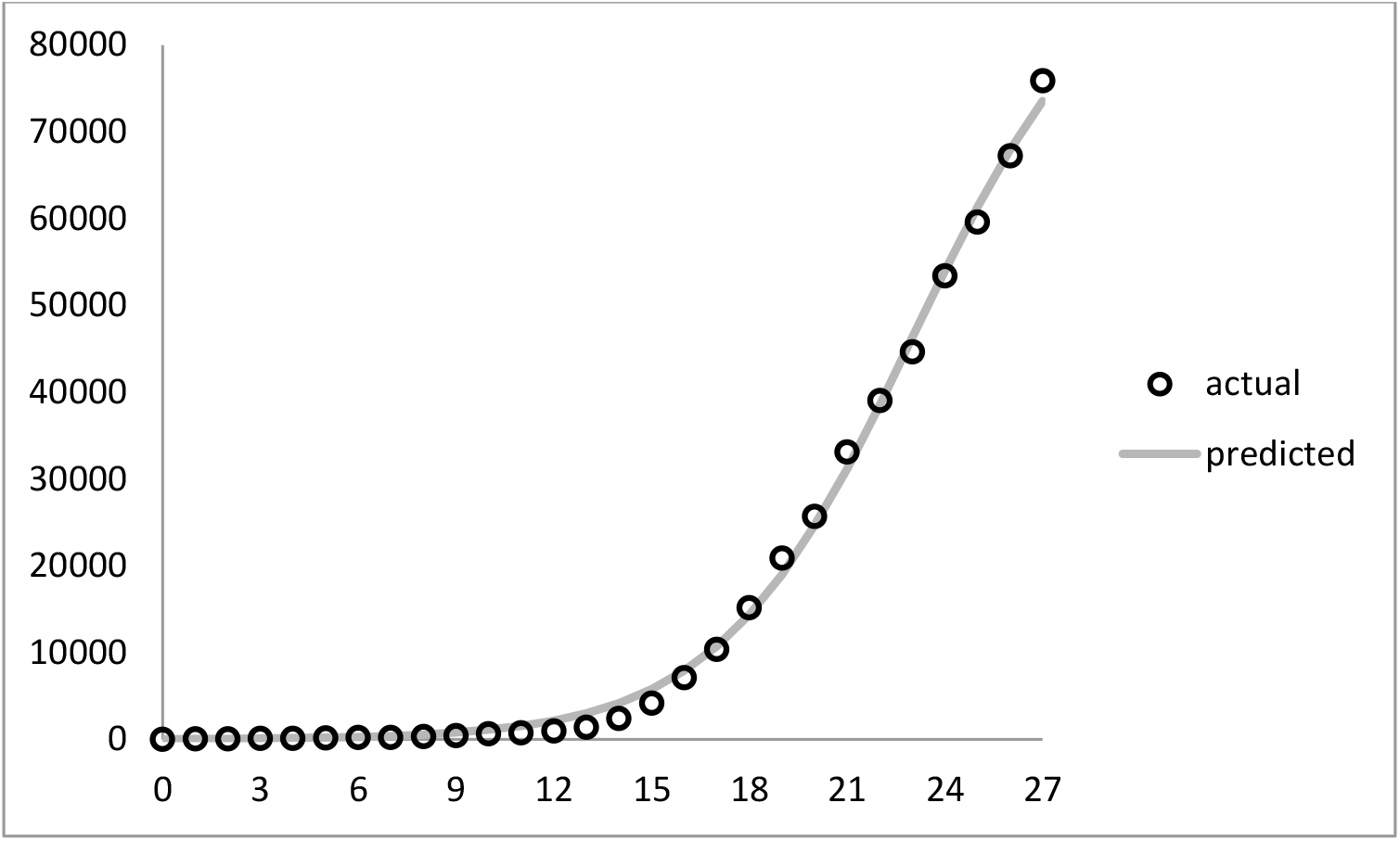
Observed cases (circles) and fitted values (continuous grey line) based on the EAP estimates for the parameters reported in Table 1. The values on the x-axis represents number of days since the initial data point used in our model (March the 4th).

Both Fig. 1 (top) and Fig. 2 show that the model’s estimates (e.g. K) are very conservative. This is a commonly observed situation for phenomenological (i.e. purely data-driven) models of this type.

At the time of writing this note, there is information for the number of infectees three days after the 28 days used in order to fit the model. We used the posterior estimates in order to predict the number of future infectees. More precisely, we used the posterior estimates (including σ) in order to simulate data for future values of t thereby constructing what is known as posterior predictive distributions. For example, for a given future day (e.g. for the 28^th^ day) we sampled all posterior values for Eq. 2 parameters and for each sample we plugged in the t=28 value in order to obtain a mean prediction value; then we added a random number generated from N(0, σ) where the value for σ is sampled from the posterior alongside the other parameters available for the given step. The resulting predictive distribution has an observed mean, variance, etc. and can be used in order to make point and/or interval predictions (HDIs) as usual. Some results are shown in Table 2.

**Table 2.** 95% and 99% HDIs for the predictive distributions for 7 days after the final data point used to fit the model. Ranges for the HDIs are given below the upper values for a given interval. The true values available by now are shown on the last row. The last column (day 200) gives the posterior prediction for the final number of infectees.

| t | 28 | 29 | 30 | 31 | 32 | 33 | 34 | 200 |
| --- | --- | --- | --- | --- | --- | --- | --- | --- |
| HDI2.5% | 74289 | 77225 | 79240 | 80764 | 81840 | 82597 | 83133 | 84323 |
| HDI97.5% | 81709 | 86016 | 89392 | 92143 | 94256 | 95883 | 97099 | 100504 |
| Range | 7420 | 8791 | 10152 | 11379 | 12416 | 13286 | 13967 | 16181 |
| HDI0.5% | 72982 | 75552 | 77644 | 79032 | 79792 | 80452 | 80896 | 81759 |
| HDI99.5% | 83071 | 87457 | 91387 | 94458 | 96663 | 98520 | 99921 | 103831 |
| Range | 10089 | 11905 | 13743 | 15426 | 16871 | 18068 | 19025 | 22072 |
| True | 83889 | 92770 | 102870 |  |  |  |  |  |

We see that the predictive distributions fail to capture even the immediate true value which once again suggests that indeed the model is inadequate and fails to capture the true trends in the data. Note, however, that the ranges of the prediction intervals increase for later data points which is a desirable quality of a model and is intrinsic to the Bayesian approach employed here. As Fig. 2 (upper right portion) suggests, the model converges too quickly to its upper asymptote and hence its predictions are too low and probably too narrow. This observation is not surprising given that it is well-known that the simple logistic model is applicable only during specific stages of an outbreak and/or when enough data is available (see Wu et al, 2020 for a review). Possible solutions include: improving the model (e.g. Richards, 1959) by adding more parameters which can account better for the deviation of the observed data points from the symmetric S-shaped curve suggested by the logistic growth model; adjusting the prior distributions so as to reflect our expectations of a much higher upper asymptote (K); switching to a different, preferably more mechanistic approach altogether.

Instead, we attempted to construct a more accurate model within the same logistic growth paradigm in a different way: we introduced weights to our data points with later data points receiving higher weights than older ones in the hope that this will alleviate some of the problems observed above. Specifically, we weighted the points according to a Rectified Linear-like function (e.g. Glorot et al., 2011) whereby the first 20 observations received constant (0.008) low weights and the last 8 observations received linearly increasing higher weights (last 8 weights=[0.77 1.55 2.32 3.09 3.87 4.64 5.41 6.19]). The idea behind this scheme was to try to force the model to account better for the observations following the approximately linear trend observed in the upper half of Fig. 2. Note also that the weights sum to the number of original observations (28). The weights pattern is shown in Fig. 3.

**Figure 3.**
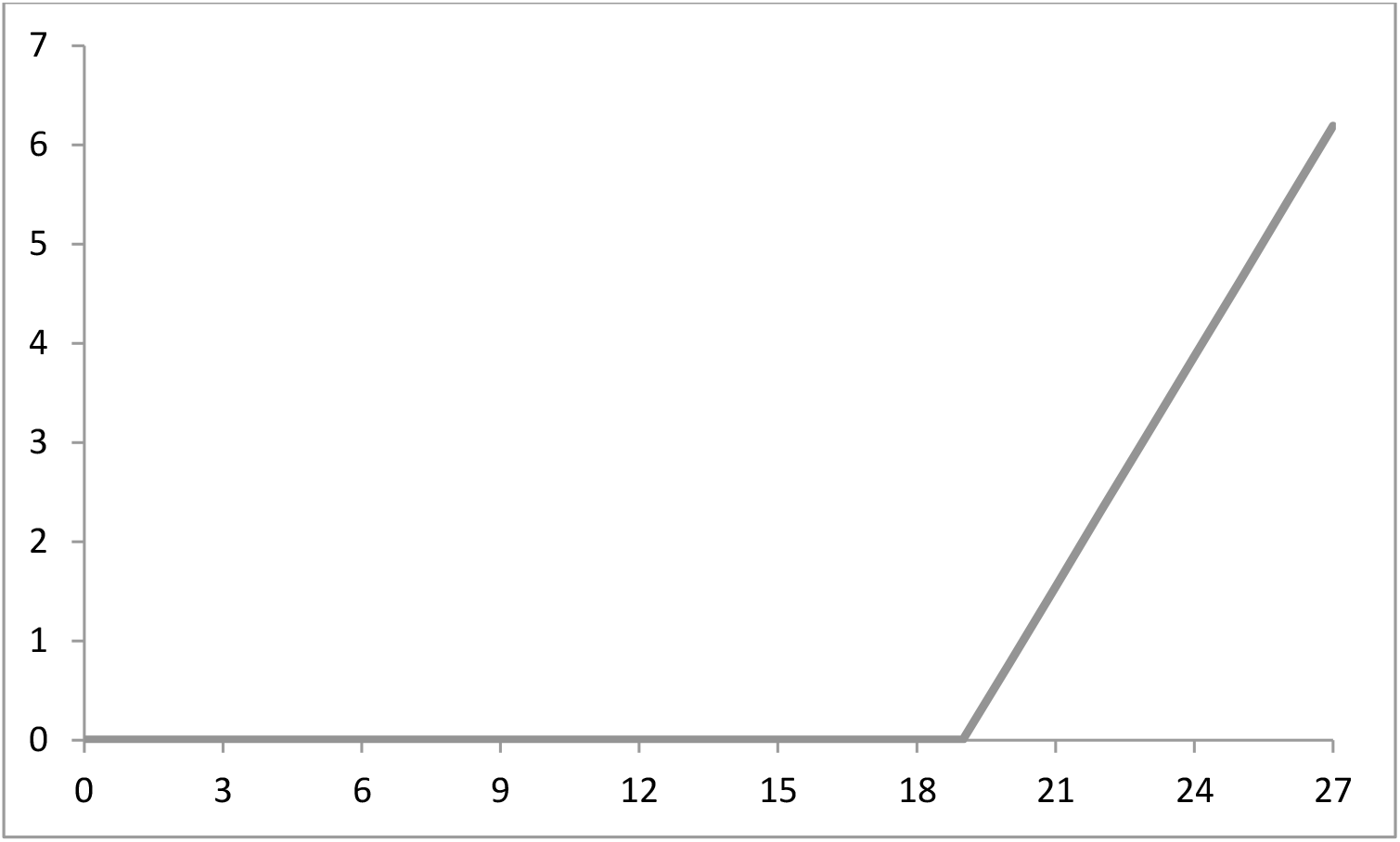
The Rectified Linear pattern of weights used to weigh the likelihood function in our second and third simulations. The x-axis denotes the consecutive number of each observation and the y-axis shows its weight. See the text for the actual values employed.

In the subsequent simulations we used the proposed weights in order to weigh the likelihood function of the model. Following Simeckova (2005), assuming we have observations Y_1_, …Y_n_ and Y_i_ has density f_i_(Y_i_|θ) where θ is the vector of parameters (see Eq. 2), we apply the weights vector **w**=[w_1_,…w_n_]. If we let l_i_(θ)=log(f_i_(Y_i_|θ)), the weighted log-likelihood function of our model becomes:

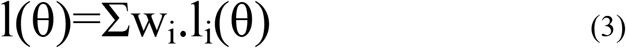

## 3. Simulation 2

We used the above weighing scheme and repeated the previous simulation. In that sense we altered the likelihood function while leaving the prior distributions intact. Everything else (including the simulation details such as number of posterior draws, thinning, etc.) was the same as reported in Section 2. Again, we observed good convergence for all parameters (see Fig. 4 depicting a traceplot and a histogram for the K parameter).

**Figure 4.**
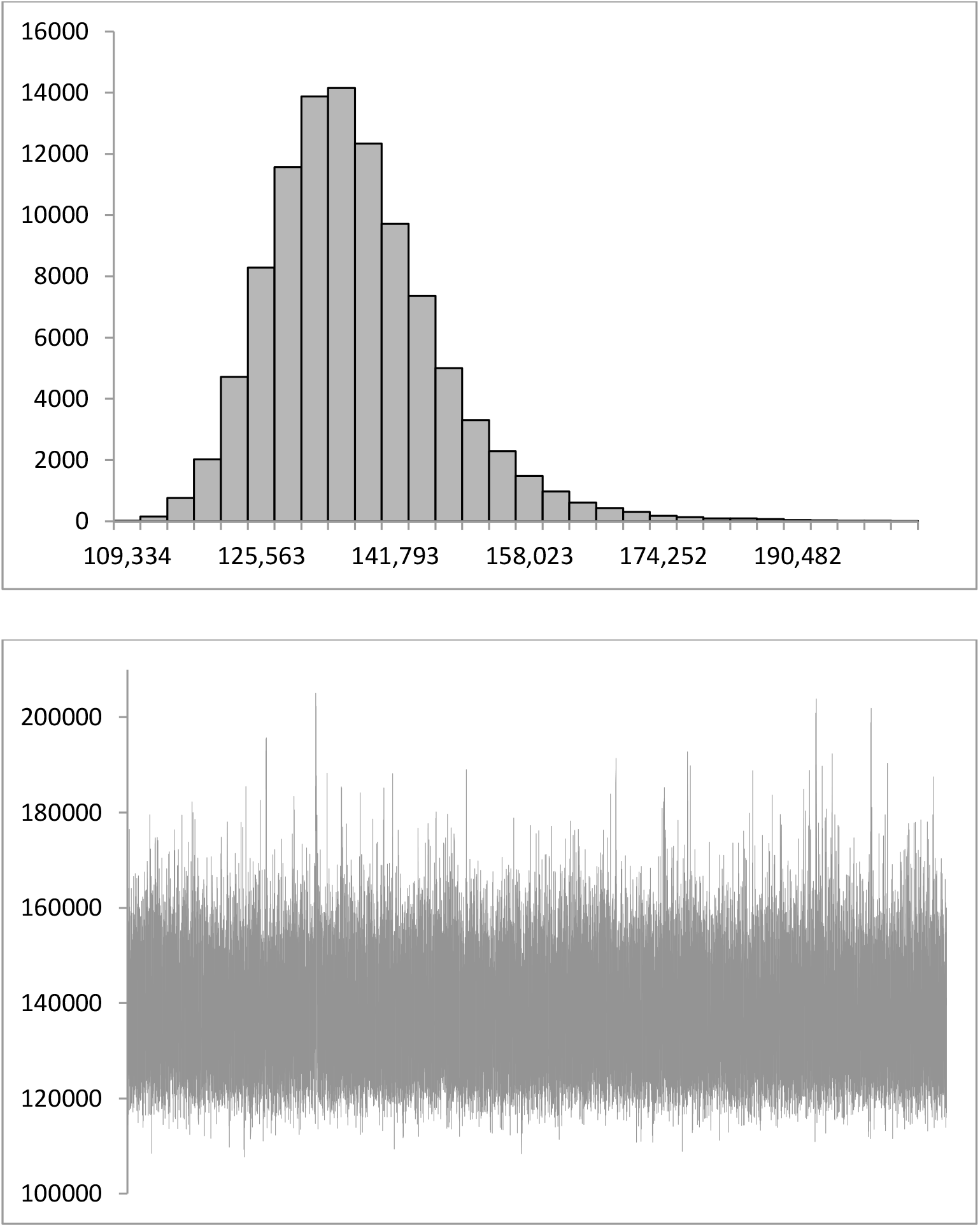
A histogram (top) and a traceplot (bottom) for the posterior for the K parameter from Eq. 2 for our second model.

Table 3 gives a summary for the posterior estimates^3^. We see that our weighting scheme appears to give more reasonable results and that the estimates for the upper asymptote (K) are substantially higher than before. The same observations can be made when we inspect the fitted equation against the observed data (Fig. 5). It is clear that the fitted curve is much more affected by the later points and consequently the upper asymptote is higher than before (compare also Tables 3 and 1).

**Table 3.** Means, medians, standard deviations (SD), and 95% HDIs for the parameters of Eq. 2 obtained from the posterior of our second (weighted) model.

|  | mean | median | SD | 2.50% | 97.50% |
| --- | --- | --- | --- | --- | --- |
| K | 136869.67 | 135614.03 | 10422.80 | 117921 | 157535.3 |
| A | 410.97 | 401.77 | 73.45 | 279.37 | 557.52 |
| r | 0.23 | 0.23 | 0.01 | 0.21 | 0.25 |
| $\sigma$ | 0.01 | 0.01 | 0.002 | 0.01 | 0.01 |

**Figure 5.**
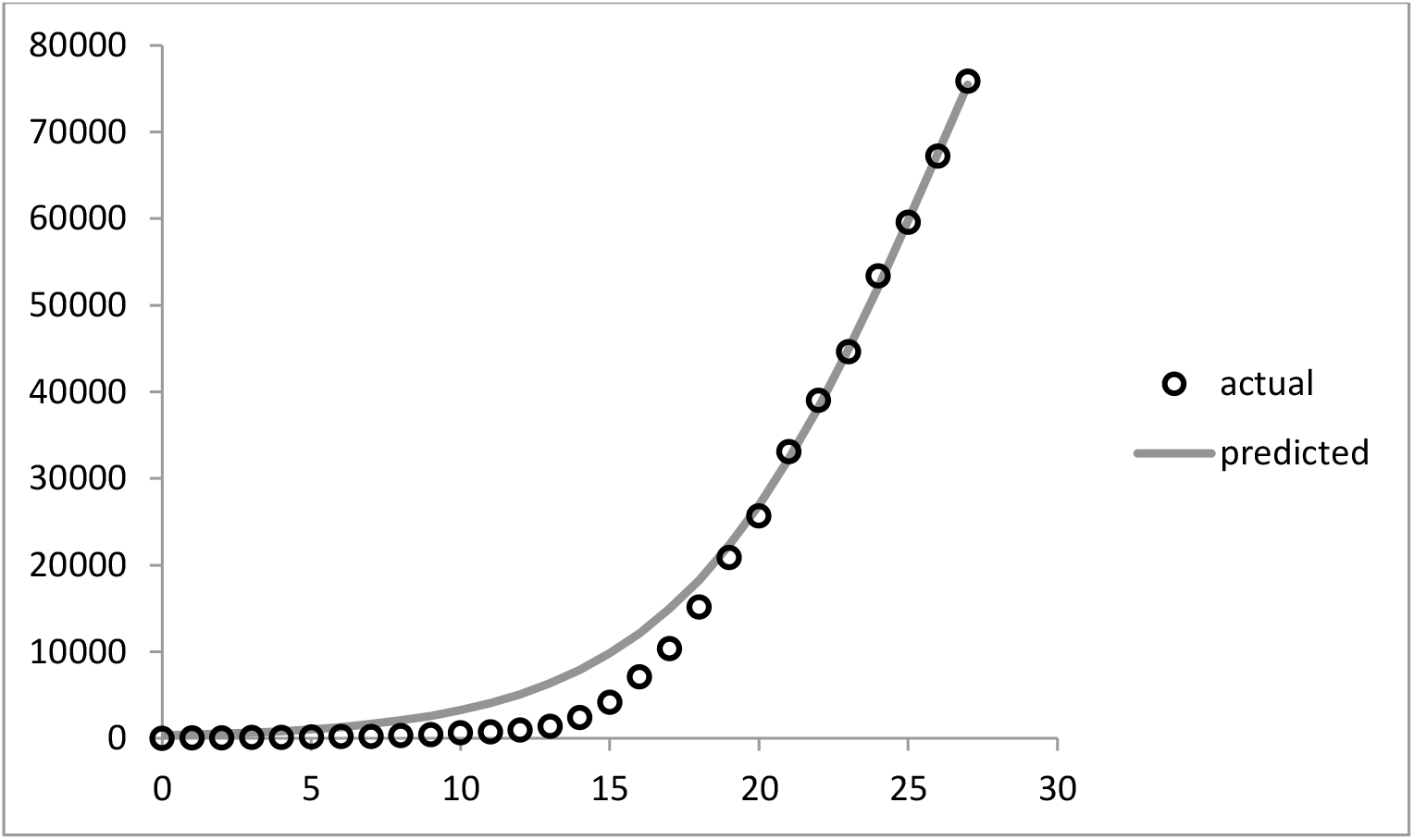
Observed cases (circles) and fitted values (continuous grey line) based on the EAP estimates for the parameters reported in Table 3.

Table 4 gives the posterior predictive distribution for 7 days ahead as well as for the estimate for the final number of infectees.

**Table 4.** 95% and 99% HDIs for the predictive distributions for 7 days after the final data point used to fit the second model. Ranges for the HDIs are given below the upper values for a given interval. The true values available by now are given at the last row (the values in bold are accurately predicted by the model). The last column (day 200) gives the posterior prediction for the final number of infectees.

| t | 28 | 29 | 30 | 31 | 32 | 33 | 34 | 200 |
| --- | --- | --- | --- | --- | --- | --- | --- | --- |
| HDI2.5% | 81208 | 87755 | 93408 | 98316 | 102300 | 105708 | 108498 | 117813 |
| HDI97.5% | 84970 | 92915 | 100644 | 108090 | 114896 | 121216 | 126905 | 157554 |
| Range | 3761 | 5160 | 7236 | 9774 | 12596 | 15508 | 18407 | 39741 |
| HDI0.5% | 80551 | 86843 | 92130 | 96700 | 100366 | 103302 | 105775 | 114050 |
| HDI99.5% | 85653 | 93872 | 102046 | 110128 | 117752 | 124759 | 131281 | 171023 |
| Range | 5102 | 7028 | 9916 | 13428 | 17386 | 21457 | 25506 | 56972 |
| True | **83889** | **92770** | 102870 |  |  |  |  |  |

We see that this time the model accurately predicts two consequent data points and fails to predict the third. This is still not a satisfactory performance, however, and hints towards the possibility that the actual process exhibits steeper rise than the one suggested by the model. In the same way, it appears that the HDIs are not wide enough in order to accommodate the actual uncertainty.

## 4. Simulation 3

Looking at figures 2 and 5 we see that the errors, in all likelihood, both lack homoscedasticity and possess an auto-correlated structure. In order to (partially) alleviate these problems we removed the normality assumption present above and replaced it with the assumption that the errors follow a t-distribution with location parameter equal to 0 and scale (similar to the standard deviation used above) and degrees of freedom (df) parameters estimated from the data (see Kruschke, 2012 for the same approach in the context of a linear model).

We used the same weighing scheme as above and introduced the two new parameters (scale and df) describing the t-distribution governing the model’s errors. We again proposed the first four parameters of the model (i.e. K, A, r, scale) by a multivariate normal distribution centered on the previous values of the chain with scaled variance-covariance matrix estimated from pilot chains; the df parameter was proposed separately based on a lognormal distribution^4^ which was transformed back to the original scale after the end of the simulation. 30 million samples from the posterior were obtained with every 300^th^ step retained (i.e. we had a thinning parameter of 300). We used improper uniform priors for all parameters except for the df parameter for which a shifted exponential with mean equal to 30 was specified as suggested by Kruschke (2012).

The results indicated good convergence (a traceplot for the K parameter is shown in Fig. 6 below; histograms from the posterior for all parameters are shown in Appendix A).

**Figure 6.**
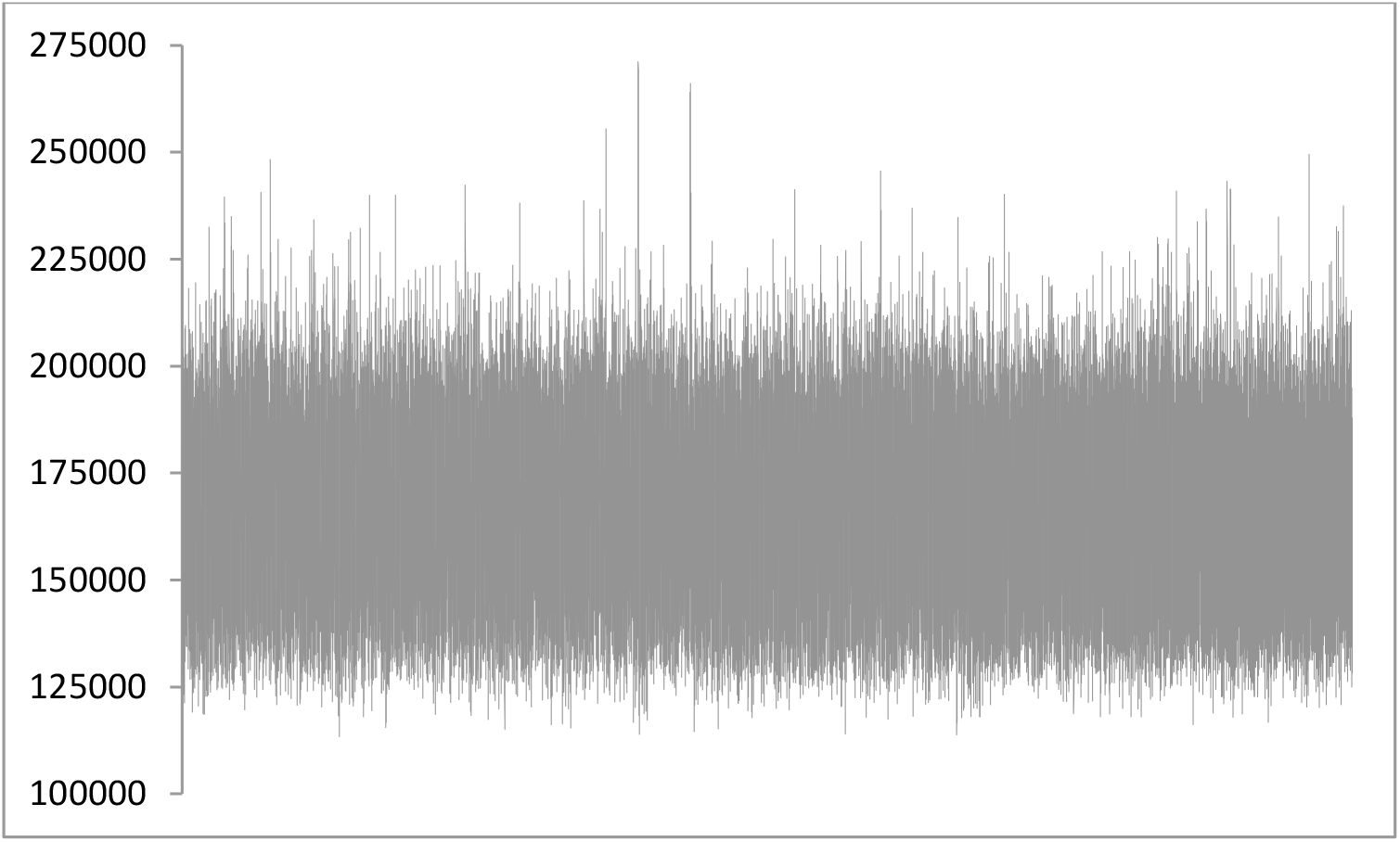
Traceplot for the posterior for the K parameter from Eq. 2 for our final model.

Table 5 gives the point and interval estimates for the posteriors for the five parameters in question. We see that the estimates differ substantially from the ones reported above and that a steeper curve is indicated.

Consistently with our expectations the 95% HDI for the df parameter suggests a noticeable deviation from normality.

Fig. 7 shows the predicted trend based on the EAP estimates shown in Table 5.

**Figure 7.**
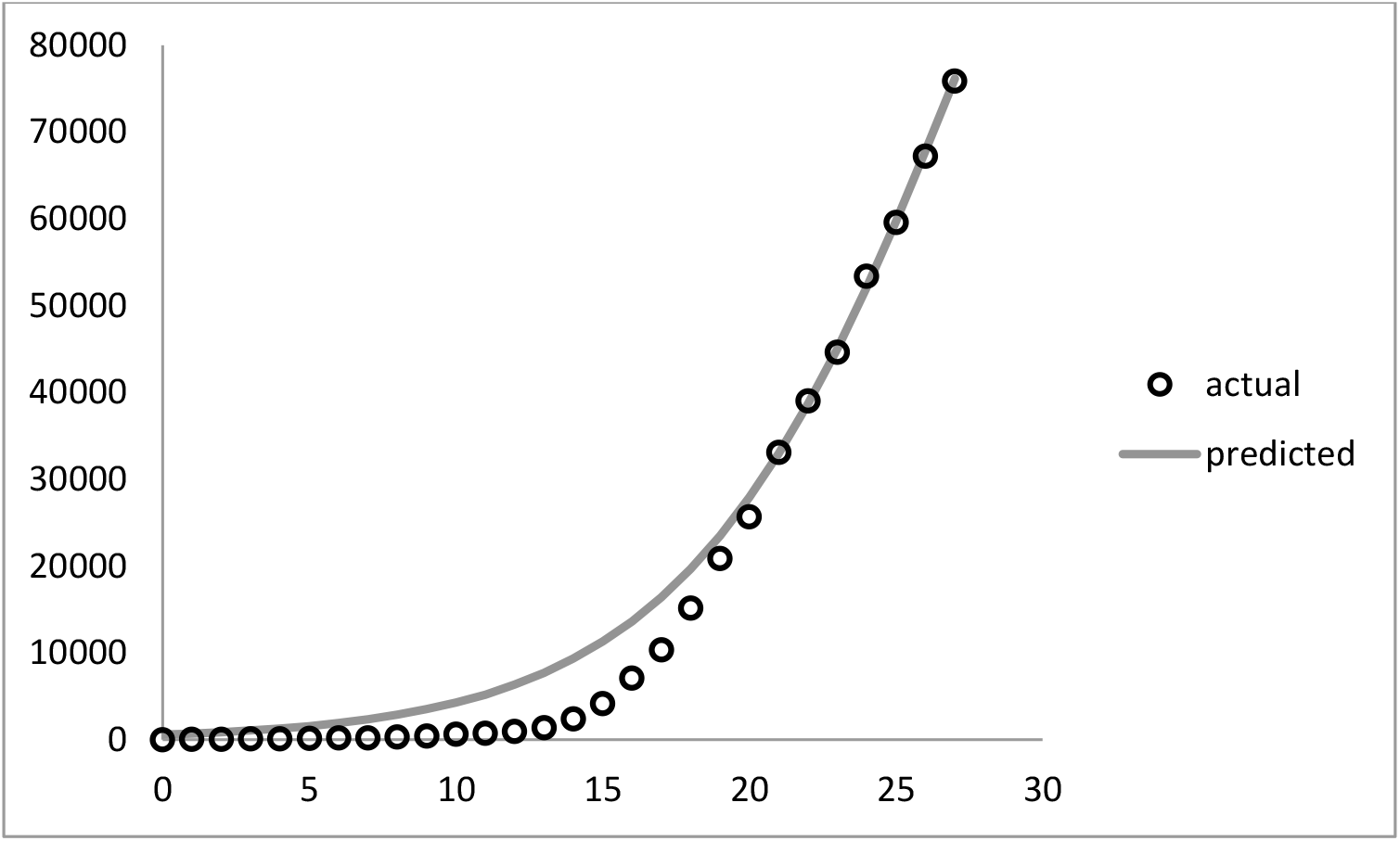
Observed cases (circles) and fitted values (continuous grey line) based on the EAP estimates for the parameters reported in Table 5.

Finally, Table 6 specifies the predictive distributions for the next 7 days and for the estimate for the final cumulative number of infectees. We see that this model accurately predicts at least three future data points. In the next several days we should be able to observe how the model deals with data points further away in time.

**Table 6.** 95% and 99% HDIs for the predictive distributions for 7 days after the final data point used to fit our last model. Ranges for the HDIs are given below the upper values for a given interval. The true values available by now are given at the last row (the values in bold are accurately predicted by the model). The last column (day 200) gives the posterior prediction for the final number of infectees.

| t | 28 | 29 | 30 | 31 | 32 | 33 | 34 | 200 |
| --- | --- | --- | --- | --- | --- | --- | --- | --- |
| HDI2.5% | 82041 | 89429 | 96073 | 101971 | 107125 | 111375 | 115128 | 129412 |
| HDI97.5% | 85694 | 94827 | 104246 | 113682 | 123002 | 131784 | 140337 | 203450 |
| Range | 3653 | 5399 | 8173 | 11711 | 15877 | 20408 | 25209 | 74038 |
| HDI0.5% | 80770 | 87838 | 94098 | 99462 | 103842 | 107458 | 110558 | 119862 |
| HDI99.5% | 87387 | 96109 | 105515 | 115211 | 124839 | 134255 | 143455 | 216170 |
| Range | 6617 | 8271 | 11417 | 15750 | 20996 | 26797 | 32897 | 96307 |
| True | **83889** | **92770** | **102870** |  |  |  |  |  |

As can be seen in Appendix A the posteriors for the different parameters no longer resemble parametric distributions (for the previous two models the posteriors for the K, A, and r definitely resemble normal/t-distributions while the σ parameter is pronouncedly positively skewed and thus resembles a gamma distribution). Nevertheless this model appears to be best suited for the modeled phenomenon.

## 5. Discussion

It appears that a logistic growth model with a weighted likelihood function and a t-distribution imposed on the error structure is able to make accurate short term predictions of the spread of a disease. The Bayesian estimation gives more accurate estimates than traditional Least Squares and Maximum Likelihood approaches with more accurate interval estimates. Moreover, the Bayesian posteriors (including the predictive distributions) have a straightforward probabilistic interpretation which cannot be said about traditional frequentist Confidence Intervals.

As a rule, the posterior distributions show high correlations between the parameters which makes algorithms like blockwise Metropolis-Hastings more effective in general than algorithms which explore a single posterior distribution at a time such as the Gibbs Sampler and the componentwise Metropolis-Hastings. The fact that some posteriors lack closed-form solutions is another impediment when it comes to the Gibbs Sampler but not necessarily for the use of the componentwise Metropolis-Hastings with respect to certain parameters as demonstrated in Section 3.

The weighing scheme employed here proves beneficial over modeling the raw data by forcing the model to pay more attention to more recent observations. Other weighing schemes are certainly possible and investigating the properties of different approaches seems a potentially fruitful future enterprise.

As a whole it appears that the combination of Bayesian Estimation, differentially weighing the observations, and employing a more robust approach towards modeling the errors (i.e. assuming a t-distribution with scale and df parameters estimated from the data) results in more reliable HDIs and prediction intervals than more traditional approaches.

Far from being perfect, the proposed model appears to be somewhat useful. Of course, such a model can be continuously augmented by including new data points and applying the same or similar weighing procedure.Presumably, continuously adjusting the model by adding new observations as they become available would improve its accuracy.

That being said, our simulations suggest that we should be somewhat skeptical towards logistic growth models applied to the raw data describing an outbreak, especially when the number of available data points is relatively small and the upper asymptote appears not to have been approached yet.

## Data Availability

All data used in this submission is freely available at the github repository. The computer codes used to implement the described calculations are provided as an Appendix.

## APPENDIX A

Histograms of the posterior distributions for the five parameters for our last model. For simplicity the scale parameter values are presented in their standardized form (i.e. they are not back-transformed).

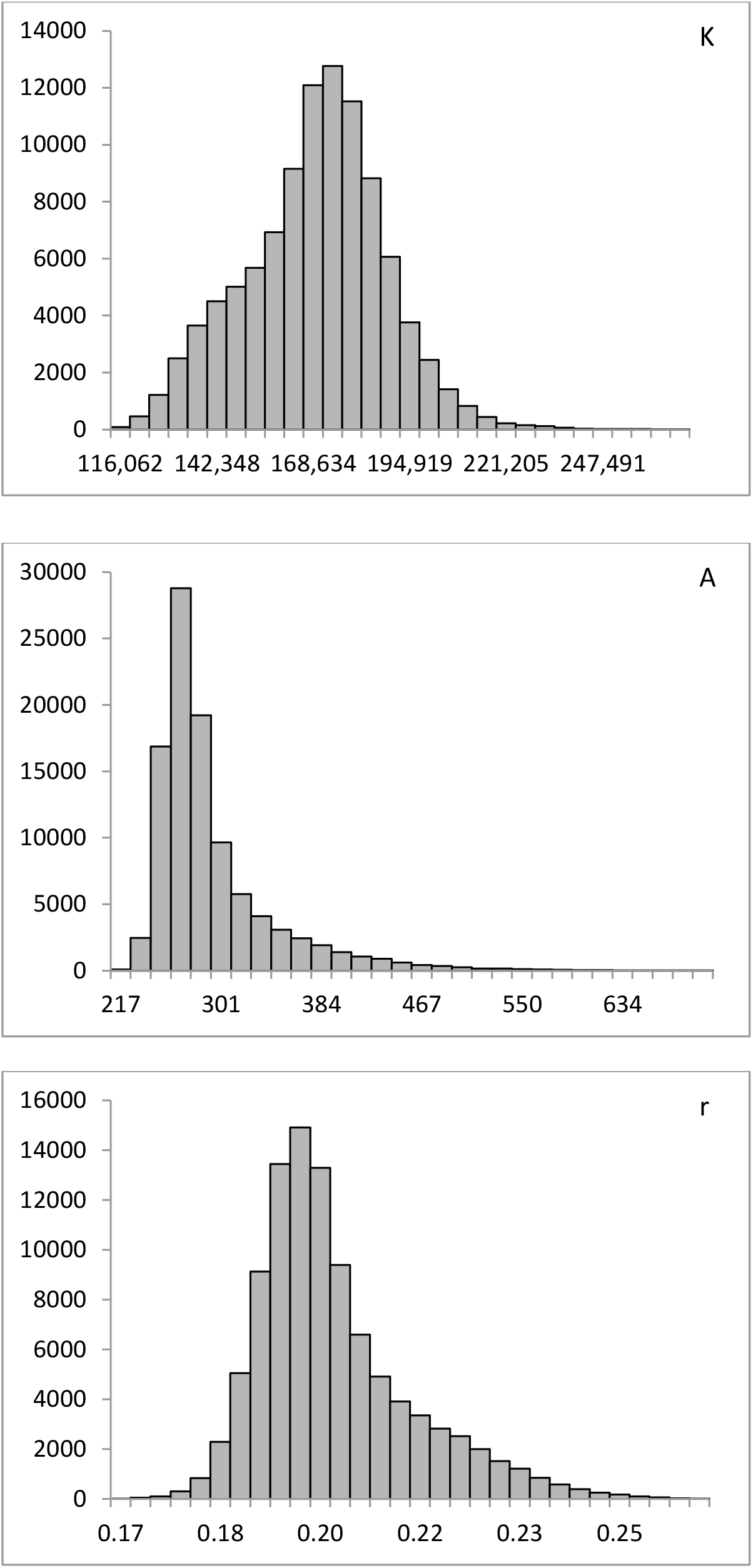
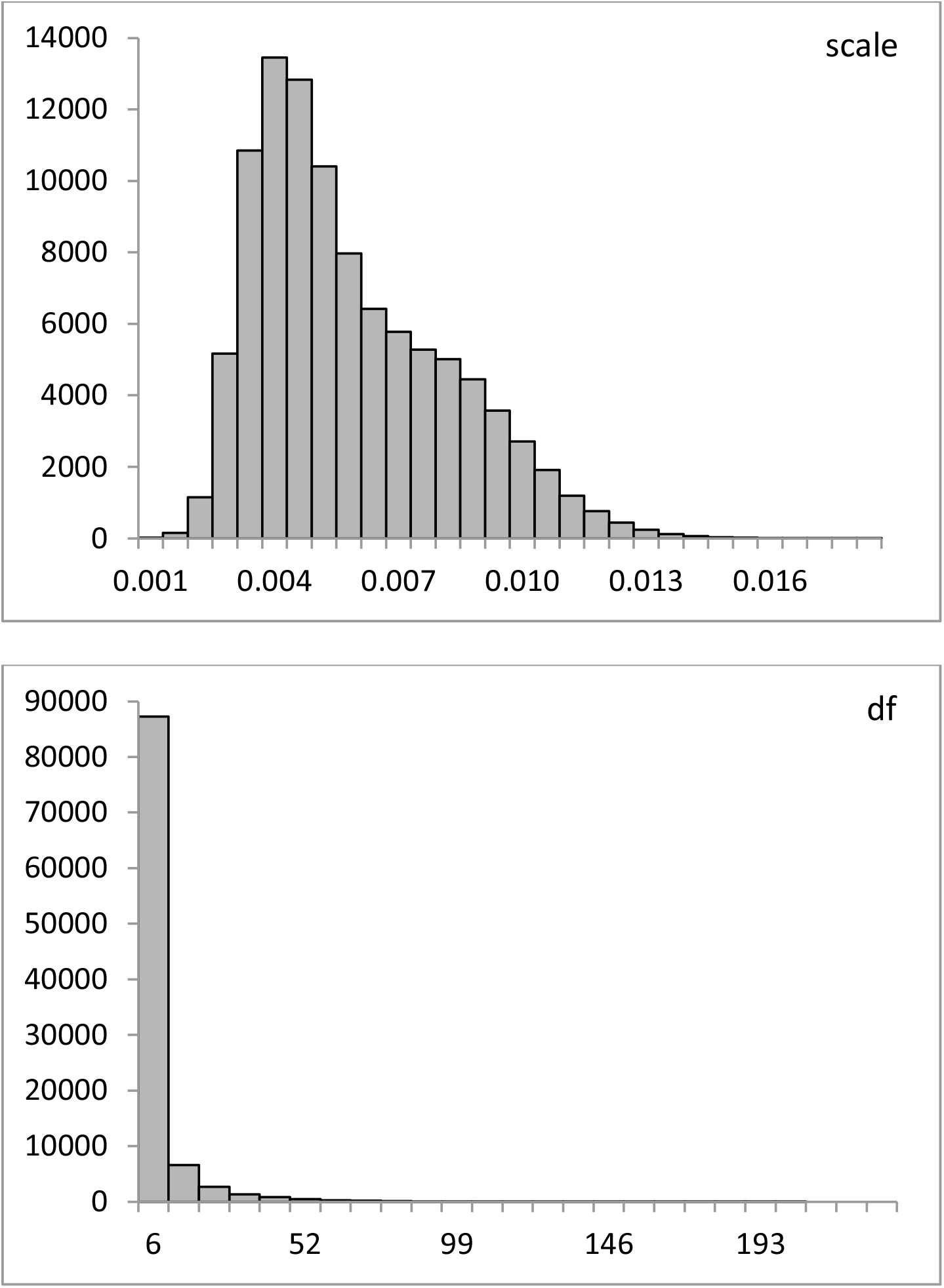

## APPENDIX B

Matlab code for the third simulation.

%t, y, and covmat should be imported before running the code; t is time in days (the first day

%for which there are 11 infectees is labeled 0 – March 4^th^); all consecutive days up to 27 are

%simply given their corresponding numbers (i.e. 0, 1, 2, 3…27); y is the cumulative number

%of infectees for each consecutive day up to the 31^st^ of March; available at github;

%the y values are standardized before the code is run by dividing them by 70000;

%The columns of PAR correspond to the posteriors for K, A, r, scale, and df respectively;

~~~
nit=30000000; %# iterations;
PAR=zeros(nit/300, 4); cur=[2.38 299 0.2046 0.0061]; %starting values; DF=zeros(nit/300, 1); curdf=0.1; %starting value for df;
wei=[0.1*ones(1, 20) [10 20 30 40 50 60 70 80]]; wei=wei/sum(wei)*28; wei=wei’; PrevPred=cur(1)./(1+cur(2)*exp(-cur(3)*t)); cnt=0;
Lp=gentdst(y, PrevPred, cur(4), exp(curdf));
Lp=sum(wei.*log(Lp)); Lp=Lp+log((1/29)*exp(-(1/29)*(exp(curdf)-1))); %posterior;
for i=1:nit
  prop=mvnrnd(cur, covmat*0.09, 1); %proposal;
  if prop(1)<=0 ‖ prop(2)<=0 ‖ prop(3)<=0 ‖ prop(4)<=0 %prior;
    alp=0;
  else
    Pred=prop(1)./(1+prop(2)*exp(-prop(3)*t));
    Ln=gentdst(y, Pred, prop(4), exp(curdf)); %Likelihood;
    Ln=sum(wei.*log(Ln)); Ln=Ln+log((1/29)*exp(-(1/29)*(exp(curdf)-1))); alp=exp(Ln-Lp); %acceptance probability;
  end
  if rand<alp
    cur=prop;
    Lp=Ln;
  end
  propdf=curdf+randn(1, 1)*0.93; %proposal for df;
  if propdf<0 %prior (df should be>=1);
    alp=-99999;
  else
    Pred=cur(1)./(1+cur(2)*exp(-cur(3)*t));
    Ln=gentdst(y, Pred, cur(4), exp(propdf));
    Ln=sum(wei.*log(Ln)); Ln=Ln+log((1/29)*exp(-(1/29)*(exp(propdf)-1))); alp=(Ln-Lp)+(log(propdf)-log(curdf)); %accounting for the log-normal proposal;
  end
  if log(rand)<alp;
    curdf=propdf;
    Lp=Ln;
  end
  if mod(i, 300)==0
    cnt=cnt+1;
    PAR(cnt, :)=cur;
    DF(cnt)=curdf;
  end
end
~~~

%%%%%%%%%%%%%%%Proposal Covariance Matrix%%%%%%%%%%%%%%%%

%should be imported as covmat to Matlab before running the above program;

%%%%%Function calculating the density for the Generalized t-distribution%%%%%%%% %the function is used to calculate the likelihood;

~~~
function y = gentdst(x, m, s, v)
%x – data point, m – location, s – scale, v 0 degrees of freedom;
c=(1/sqrt(v))*(1/(beta(v/2, 0.5)));
y=(c/s)*(1+((x-m).^2)/(v*(s^2))).^(-0.5*(v+1));
~~~

|  | K | A | r | scl |
| --- | --- | --- | --- | --- |
| K | 0.07903 | -12.97633 | -0.00380 | -0.00047 |
| A | -12.97633 | 2869.77687 | 0.71946 | 0.08648 |
| r | -0.00380 | 0.71946 | 0.00020 | 0.00002 |
| scl | -0.00047 | 0.08648 | 0.00002 | 0.00001 |

## Footnotes

1 The data is publicly available via the github repository: https://github.com/nytimes/covid-19-data/blob/master/us-states.csv

2 The posterior chains showed high correlations between the parameters (ranging from -0.87 to +0.96) which renders algorithms like the Gibbs Sampler and/or componentwise Metropolis highly inefficient for the task at hand.

3 For comparison the LS estimates obtained with the same weighing scheme are K=135870 (95% CI=[117600; 151140]), A=398.5 (95% CI=[267; 530.1]), and r=0.23 (95% CI=0.21; 0.25]).

4 The lognormal distribution is not symmetric and hence we use the actual Metropolis-Hastings acceptance probability (e.g. Chib & Greenberg, 1995) during the step sampling from the df posterior.

## References

Batista, M. (2020). Estimation of the Final Size of Coronavirus Epidemic by the Logistic Model. medRxiv, 2020.

Chib, D., Greenberg, E. (1995). Understanding the Metropolis-Hastings Algorithm. The American Statistician, 49, 327–335.

Glorot, X., Bordes, A., Bengio, Y. (2011). Deep Sparse Rectifier Neural Networks. Proceedings of the 14th International Conference on Artificial Intelligence and Statistics, FL, USA, Vol. 15.

Kruschke, J. (2012). Bayesian Estimation Supersedes the t test. Journal of Experimental Psychology: General, 142 (2), 573–603.

Richards, F. (1959). A Flexible Growth Function for Empirical Use. Journal of Experimental Botany, 10 (29), 290–301.

Simeckova, M. (2005). Maximum Weighted Likelihood Estimation in Logistic Regression. WDS’05 Proceedings of Contributed Papers, 1, 144–148.

Wu, K., Darcet, D., Wang, W., Sornette, D. (2020). Generalized Logistic Growth Modeling of the COVID-19 Outbreak in 29 Provinces in China and in the rest of the World. arXiv, 2020.

